# Keep it CooL! Results of a two-year CooL-intervention: a descriptive case series study

**DOI:** 10.1101/2023.06.15.23291479

**Authors:** Nicole Philippens, Ester Janssen, Stef Kremers, Rik Crutzen

## Abstract

**Background:** Coaching on Lifestyle (CooL) is a two-year healthcare intervention for people with overweight or obesity, stimulating weight reduction by promoting sustained healthier behavior. The objective of this study is to investigate the effects of CooL on participants’ anthropometrics, personal factors and behavioral factors over the two-year timeframe of CooL.

**Methods:** A descriptive case series study, using a broad set of routinely collected data on anthropometrics, personal factors and behavioral factors of adults living across the Netherlands. The data were collected between November 2018 and December 2021 among participants of CooL (N=746) at three moments during the intervention: at baseline (T0), at 8 months (T1) after completion of phase 1 and at 24 months (T2) after ending CooL. Changes over time were analyzed using paired t-tests comparing baseline to T1 and baseline to T2. In addition, potential differences on outcomes in subgroups based on education level, weight status and group size were examined using paired t-tests and ANOVA-tests.

**Results:** The results showed changes in the desired direction on all outcomes at 24 months compared to baseline. The largest effects were seen on perceived health, attentiveness towards meal size and meal composition (large effect size). Mean weight loss was 4.13 kilograms (SD 7.54), and mean waist circumference decreased with 4.37 centimeters (SD 8.59), indicating a medium to large effect size. Changes in outcomes were similar across all subgroups consisting of participants with different educational level, BMI at baseline and in different group sizes.

**Conclusion:** The study showed sustained effects in weight-related outcomes of CooL over the timeframe of 24 months supporting the two-year set-up of CooL. The outcomes indicate that CooL is appropriate and effective for different group sizes and for a wide variety of participants irrespective of gender, age, BMI at baseline or level of education.

## Introduction

Obesity is considered a chronic disease according to the World Health Organisation [1] and the Dutch Health council [2] and it is linked to many other diseases – both physical and mental [3] – and a diminished quality of life [4].

Overweight is more common among men (53%) than women (47%), obesity on the other hand is more prevalent in women (17%) than in men (12%). Approximately 41% of people with a higher level of education are overweight whereas this percentages rises to 60% for people with a lower level of education. The proportion of people with severe obesity is twice as high among people with a lower level of education (18%) compared to those with a higher level of education (10%) [5].

Consensus has been reached internationally on the importance of an integrated approach to target overweight and obesity, including limited energy intake, healthy food choices and regular physical activity [6]. The Dutch national guidelines have added stress management and sleep as additional essential elements to tackle overweight and obesity [7].

As of January 2019, Combined Lifestyle Interventions (CLIs) are part of basic health insurance. Having a basic health insurance is a legal obligation for every person living or working in the Netherlands, and as such a CLI is available for all adults meeting the inclusion criteria (i.e. being obese (BMI>30) or being overweight (25<BMI<30) combined with comorbidity; and being sufficiently motivated).

Coaching on Lifestyle (CooL) is one of these CLIs; a two-year healthcare intervention for people with overweight or obesity, stimulating weight reduction by promoting sustained healthier behavior. The set-up of CooL is in line with the recommendations of the WHO. In addition, the outcomes of the first eight months of CooL, even during COVID-19 and its accompanying restrictions, showed substantial and promising results. Both aspects make CooL an appropriate intervention for people that are overweight or obese [6, 8].

Research on long term effects of lifestyle and/or behavior change interventions has been done mainly on specific patient groups and disease related outcome measures [9]. The long-term effects of CLIs (including CooL) in the Netherlands are still unknown, mainly due to the short timespan that the CLI is currently running. So far, effect outcomes over the full intervention period are limitedly available and when outcomes are available, they are showing a stabilization or small relapse in the second year of the intervention [10]. This is the first research on the changes over time on participants over the full CooL-intervention course of 24 months.

We suspect the changes in outcomes to vary with differences in participants and with differences in the context in which the intervention is executed. As CooL aims at higher levels of self-management and self-steering we suspect that the intervention is less effective for people with a lower level of education in line with previous research indicating that level of education shows a strong and positive correlation with health and health related behaviors [11, 12]. A lower BMI at baseline has previously been associated with better program adherence [13] and a higher BMI is associated with unhealthier food choices, i.e. less fruit and vegetable, less fiber, and more fried food consumption [14] whereas on the other hand health interventions seem equally appropriate for different BMI-categories at baseline [15].

These contradictory findings sparked our interest to investigate the relationship between BMI at baseline and differences in effect sizes on the outcomes of CooL as well. Furthermore, we are interested in the differences in effect of a large versus a small group size on the CooL outcomes. No consensus has yet been reached on the optimal group size for group interventions, while in CooL these group sizes vary per context. Research in education has shown that a group size of five, compared to fifteen members, enhances participation and satisfaction of the group members [16] whereas groups of nine or more participants bring diversity of thought, experiences, and viewpoints, thereby stimulating active participation of group members [17]. Group lifestyle interventions are usually offered in groups of 10 up to 15 participants [18, 19]. These mixed findings do not provide a clear picture on the optimal group size for health interventions. In our definition, aligned with the practice of CooL, large groups consist of 10 or more participants whereas small groups have less than 10 participants. We hypothesize that participants in small CooL groups show larger effect sizes on the outcomes of CooL because a smaller group provides the coach with more time and focus per participant, thereby stimulating active participation and behavior change.

Our objective is to study the effects of CooL after 24 months on anthropometrics, personal factors and behavioral factors of the participants. In addition, we want to analyze potential differences on the outcomes for people with lower education compared to medium or higher education, people with a lower BMI at baseline compared to a higher BMI and for people participating in CooL in groups under or over 10 participants.

## Materials and Methods

### CooL-intervention

CooL is a Combined Lifestyle Intervention (CLI) including a one-hour intake and phase 1 (8 months) in which behavioral change is initiated followed by phase 2 (16 months) in which both behavioral change and behavioral maintenance are targeted. The intervention consists of individual sessions (6 hours in total) and 8 group sessions (1,5 hours each) both in phase 1 and phase 2, resulting in a higher density of sessions in phase 1 compared to phase 2. CooL aims at changes in anthropometrics (i.e. weight, BMI and waist circumference) and at an increase in perceived quality of life by stimulating healthier eating habits, less sitting time, more physical activity and attention for sufficient relaxation and high quality sleep.

CooL is an open CLI, i.e. an intervention without a strict protocol. Coaches may adapt the intervention to the target group and context as long as the main effective elements of CooL (e.g., goal setting, mobilizing social support, positive psychology, self-management and self-monitoring) are respected in implementation. The CooL-coach is a trained and licensed professional who coaches participants towards a predefined set of final objectives on health-related skills and knowledge. Participants are stimulated to take responsibility for their personal lifestyle changes by addressing motivation, personal objectives and behavioral changes. The main objective is to coach and activate participants to a sustained healthier lifestyle in line with their individual needs and personal goals.

### Study design and population

As CooL is part of regular health care in the Netherlands, a control group receiving no treatment would be both unethical and impractical, making a descriptive case series study the appropriate study design in the Dutch context. The participants are adults living throughout the Netherlands. All participants met the inclusion criteria for participating in a CLI and were referred to CooL by their general practitioner, practice nurse or internist. The decision on a proper fit for inclusion was up to the participant, the referrer and the coach. All participants signed an informed consent regarding data collection for this study.

### Data collection

We used a lifestyle questionnaire and anthropometric measurements to collect a broad set of data. The lifestyle questionnaire was based on existing validated questionnaires. The outcome measures can be divided into the categories: anthropometrics (i.e. weight/BMI and waist circumference), control and support (i.e. self-mastery and social support), physical activity (i.e. sedentary time and active minutes), diet attentiveness, alcohol use and smoking, perceived fitness (i.e. perceived health, fitness and impact of stress on daily functioning), sleep and stress.

During the course of the study, the questionnaire was extended with additional questions covering changes in context (e.g. COVID-19) and adjusted with textual simplifications in both questions and answers preserving the original essence as much as possible.

### Datasets

Data were collected from November 2018 until December 2021 at three moments in time: at baseline, during the intake (T0), after completion of phase 1 (T1) and after ending CooL (T2). No information is available on the exact number of participants starting with or dropping out of the intervention as data is submitted only by participants who agreed to share data. In addition, data collection is restricted to two moments during the intervention: after 8 months (at T1) and when ending CooL (at T2). The data at T2 contains participants that completed the intervention (sent in after approximately 24 months) and participants that dropped out earlier in time (sent in at the moment of dropout, which could be at any moment during the two-year intervention). Participants with a T2 measurement were included in the dataset if their (estimated) completion date of CooL was or would have been before the end of the data collection period, i.e. December 31^st^ 2021.

All analyses were performed between May 2022 and May 2023 and were done on the full dataset (A) consisting of program finishers and dropouts, to provide a realistic reflection of the potential intervention effects in practice. In addition, we analyzed a cleaned dataset (B) including all participants that completely finished the two-year intervention, to portray efficacy of the intervention. Changes over time were measured from baseline to T1 and from baseline to T2.

#### Demographics

At baseline, participants reported their personal characteristics such as gender, date and country of birth, highest completed education, marital status, living situation and occupational status. Educational level was categorized in line with the Dutch Central Bureau of Statistics (CBS) into low (i.e., no education or primary education), intermediate (e.g., secondary education) and high (e.g., tertiary education). The living situation was divided into living together with someone (married or cohabiting) with or without kids and living alone (divorced, unmarried or widowed) with or without kids. The occupational status was categorized into working (e.g. paid work, voluntary work or self-employed) and not working (e.g. stay-at-home, unemployed, retired or student). Country of birth was categorized into Dutch or non-Dutch.

#### Anthropometrics

Under normal conditions anthropometric data (weight, length and waist circumference) were measured by the CooL-coaches with professional equipment according to the guidelines provided by the Dutch Association of General Practitioners (Dutch: Nederlands Huisartsen Genootschap, NHG) [20]. Body weight (kg) was measured in kilogram, rounded off the nearest decimal. Height (m) was measured to the nearest centimeter without shoes. Waist circumference measurements were obtained to the nearest centimeter with a tape measure. As COVID-19 restrictions could have changed the measurement method, additional information, gathered from the CooL-coaches that were the main data suppliers (representing data of 227 participants), confirmed that in general, physical measurements took place either by the coach or on a distance of 1.5 meters under direct supervision of the coach.

#### Control and support

The self-mastery questions in the questionnaire were based on the short version of the Pearlin Mastery Scale using four questions (for example “I have little control over the things that happen to me”) and a 5-point Likert scale ranging from strongly agree (1) to strongly disagree (5) [21]. To identify social support, we questioned the perceived support of close ones using a 5-point Likert scale ranging from no support at all (1) to a lot of support (5).

#### Physical activity

The outcome measurements on physical activity, diet and personal factors were defined in cooperation with the Dutch Association of Lifestyle Coaches (BLCN) with the objective to capture the essence and map the desired outcomes of lifestyle coaching in a minimum set of questions. Physical activity used questions on sedentary behavior, both on most and least active days (“What is the average number of hours you spent sitting on the day of the week you sit the most?”) and the number of physical activity minutes per day (“What is the average minutes per day that you are physically active (in minimum bouts of 10 minutes)?”).

#### Diet attentiveness, alcohol and smoking

We defined questions on dietary attentiveness, in line with the input of the BLCN, based on the idea that deliberate behavior changes start with being aware of one’s own behavior. We used questions on the attentiveness of participants towards meal composition and meal quantities and attentiveness during the actual consumption of food using a 5-point Likert scale from very little attention (1) to a lot of attention (5). At T1 and T2 an additional question was added regarding changes in eating pattern: a reflection of the perception on healthy diet improvements compared to baseline (“How much healthier have you been eating since the intake of this program?”) with the answers ranging from much healthier (1) to much unhealthier (5). The amount of alcohol and smoking was questioned by numerical values.

#### Perceived fitness

Perceived fitness existed of questions, in line with the input of the BLCN, on perceived fitness when waking up and during the day, the impact of stress on daily functioning and on perceived health (i.e. feeling good about oneself, the extent of self-care invested and the perception of one’s general health). Questions were answered using a 5-point Likert scale, ranging from not good at all (1) to very good (5).

#### Sleep

We defined a specific set of questions around the sub-constructs: subjective sleep quality, sleep latency, sleep duration, habitual sleep efficiency, sleep disturbances, use of sleep medication and daytime dysfunction, analogous to the validated and widely used PSQI- questionnaire [22]. Each subconstruct was covered by one or two question(s) using a numerical value or a 4-point Likert scale, ranging from ‘never’ (1) to ‘three times per week or more frequently’ (4).

#### Stress

For stress, the validated Perceived Stress Scale questionnaire was used, which exists of ten questions using a 5-point Likert scale from never (1) to always (5) [23].

### Statistical analyses

#### Data preparation

We recoded some of the variables to facilitate interpretation in the sense that a higher/positive score refers to a desirable trend and a lower/negative score to an undesirable trend in the variable. For constructs based on validated questionnaires (i.e. sleep and stress) we adopted the accompanying approach without recoding. Secondly, we performed an exploratory factor analysis and calculated McDonald’s omega to assess the internal structure of items regarding several constructs such as perceived health, self-mastery, sleep and stress in line with Crutzen et al. [24]. These analyses justified summarizing all lifestyle constructs by item score means. Missing data were excluded from the statistical analyses.

#### Effect sizes

For all items and constructs, we ran descriptive statistics. Changes over time were analyzed using paired t-tests comparing baseline to T1 and baseline to T2. Effect sizes were calculated and interpreted in accordance with Lipsey’s guidelines for each pair of items or constructs, i.e. an effect size smaller than 0.32 is considered small, an effect size between 0.33 and 0.55 is considered medium and an effect size above 0.56 is considered large [25]. To improve comprehensibility effect sizes were represented such that positive values represented change in the desired direction whereas negative values represented change in an undesired direction. All T-tests were performed using SPSS-software (version 27). Missing data were excluded from the statistical analyses.

To be considered successful the target for the CLI (including CooL) is an average 5% weight loss for all participants, as set by the Dutch Partnership Overweight (Dutch: PON), an advisory body for the Dutch government on obesity related health issues. We categorized the outcomes on weight: 5% weight loss or more, between 0 and 5% weight loss, weight stabilization and weight gain, to map the percentage of participants that comply with this target.

#### Subgroup analyses

We compared different subgroups in sequence to explore potential differences in outcomes, i.e. subgroups based on educational level of the participants, on BMI at baseline and on group size at the start of CooL. Subgroup analyses were done on the full dataset (A) including program finishers and dropouts. To enable subgroup comparison, we calculated the difference (delta) between T0 and T2 for each construct or variable. As a higher starting weight usually requires less effort to lose a certain amount of weight, we looked at relative (%) weight loss compared to baseline for the BMI-subgroup comparison. For the construct ‘eating pattern’ we used the construct itself as it already includes changes in eating pattern compared to baseline in the formulation of the questions. When comparing two subgroups we performed independent t-tests comparing all delta-variables. In case of multiple subgroups, we ran an ANOVA test on the delta-variables followed by post-hoc Tukey tests to analyze potential differences in effect.

#### Drop-outs

We used logistic regression analysis to determine the main factors related to drop-out. The predictor variables in the logistic regression were based on the pre-defined subgroups of interest (i.e. based on group size, BMI at baseline and educational level) and two additional demographic variables (i.e. age category and gender). For the dropout analysis we used the full dataset (A) excluding the participants (n=22) that could not be assigned as program finisher or dropout due to missing information.

The dropout analysis showed no distinct pattern in dropout profiles. However, specific subcategories of some of the constructs were less likely to drop out in comparison to the reference category, i.e. a BMI of 35-40 compared to BMI<30, participating in a group of over 10 compared to less than 10 and a higher level of education compared to a lower level of education all were less likely to drop out. The constructs gender and age showed no differences in dropout. See Appendix 1 for the details on dropout percentages and related analyses.

As CooL is part of basic health insurance and data is gathered from all participants, provided that they gave written consent for the use of their anonymized data, selection bias is limited. In addition, we tried to minimize bias by ensuring a check on all analyses by a second researcher, by including both program finishers and dropouts in our analyses and by presenting a complete set of outcomes on all variables and analyses.

#### Ethics

This study was submitted to and approved by the Research Ethics Committee of the Faculty of Health, Medicine and Life Sciences of Maastricht University (FHML-REC/2019/073). All participants gave their written informed consent for their anonymised personal data to be used for research purposes.

## Results

### Datasets

We collected data from in total 3780 participants that started CooL between November 2018 and December 2021.See Figure 1 for a graphical representation of the dataset selection steps.

**Figure 1.**
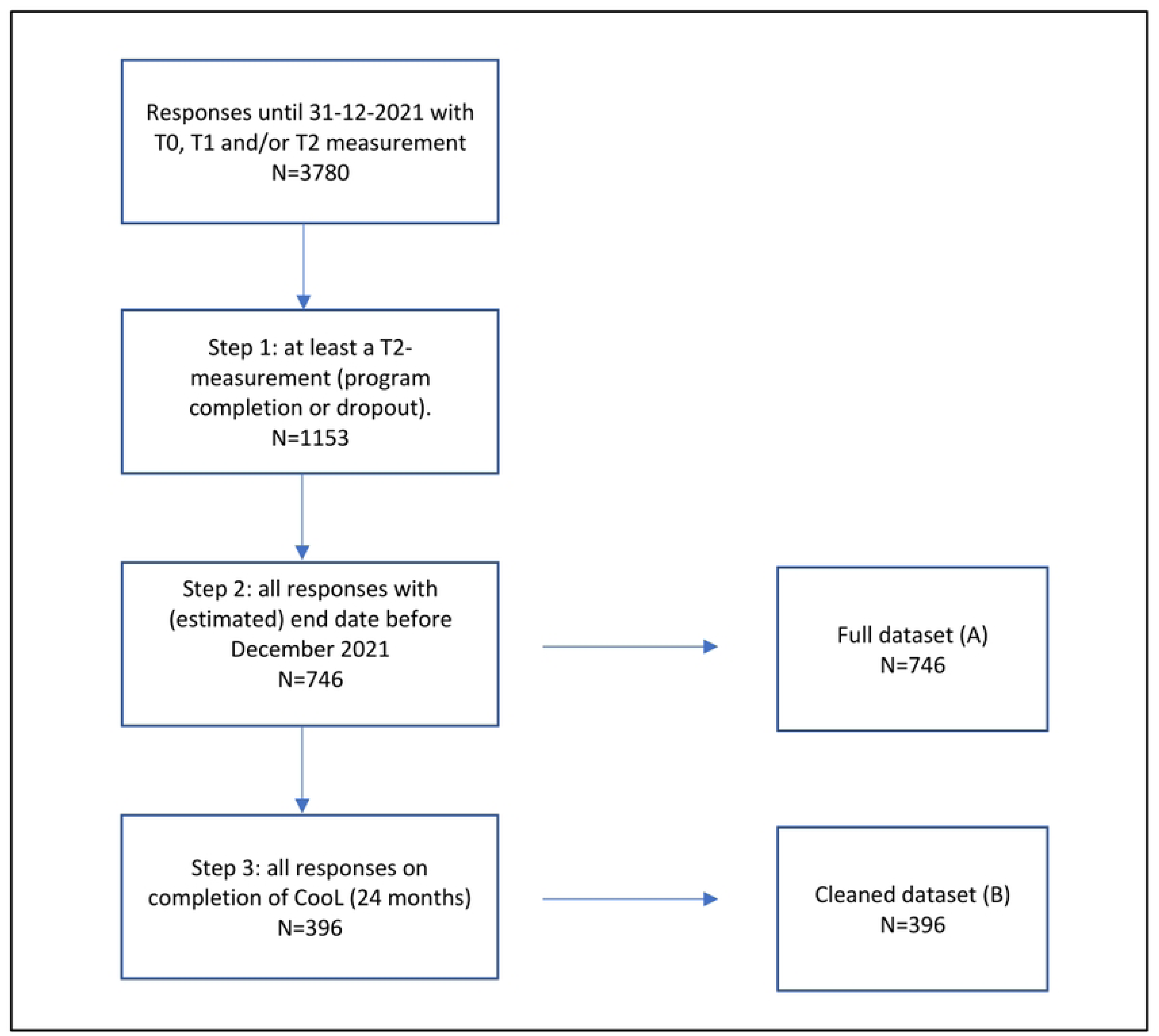
Flowchart dataset selection process.

#### Demographics

Of all participants in the full dataset (A) a total of 28% were male and 72% female. This ratio is in line with the data from the national CLI-monitor [26]. Most participants (93%) had a Dutch background. In total, 69% of the participants had a lower or intermediate level of education; 30% did not have a steady job (anymore) and approximately two third of the participants were living together with a partner (see Table 1).

**Table 1.** Demographics of the participants

| Category | Demographic | Number of participants (%)<br>full dataset (A) | Number of participants (%)<br>cleaned dataset (B) |
| --- | --- | --- | --- |
| Gender | Male | 203 (28%) | 105 (28%) |
|  | Female | 519 (72%) | 272 (72%) |
|  | Other | 1 (0.1%) | 1 (0.3%) |
| Age | Until 35 years | 75 (11%) | 41 (12%) |
|  | 35 – 44 years | 90 (13%) | 42 (12%) |
|  | 45- 54 years | 166 (25%) | 89 (26%) |
|  | 55 – 64 years | 198 (29%) | 103 (30%) |
|  | 65+ | 147 (22%) | 70 (20%) |
| Living situation | Single | 157 (22%) | 85 (22%) |
|  | Single parent | 48 (7%) | 30 (8%) |
|  | Living together with kids | 284 (39%) | 150 (40%) |
|  | Living together without kids | 200 (28%) | 98 (26%) |
|  | Other | 33 (4%) | 15 (4%) |
| Country of birth | Dutch | 668 (93%) | 348 (93%) |
|  | Non-Dutch | 49 (7%) | 27 (7%) |
| Working situation | Employed | 515 (70%) | 276 (71%) |
|  | Unemployed | 217 (30%) | 111 (29%) |
| Education | Lower level | 197 (28%) | 88 (23%) |
|  | Intermediate level | 293 (41%) | 158 (42%) |
|  | Higher level | 223 (31%) | 129 (35%) |
| **Participants** | **Total number** | **746** | **396** |

The cleaned dataset (B), containing only respondents that finished the intervention, showed in general a similar demographic picture except for the educational level of the participants: this dataset contained relatively more participants with a higher-level education and less participants with a lower-level education.

All results on the anthropometric and personal factors in the full dataset (A) are summarized in Table 2 whereas more detailed information is provided in appendix II.

**Table 2.** Overview of changes over time in anthropometrics and personal factors in complete population and in subgroups

| Changes over time on full dataset (A) at T1 and T2 | | | | | | | | ΔT0T2 comparing subgroups | | |
| --- | --- | --- | --- | --- | --- | --- | --- | --- | --- | --- |
| Construct/<br>factor | T0 M (SD) | T1 M (SD) | T2 M (SD) | $\Delta$ T0T1 [95%<br>CI] | Cohen's<br>d**<br>T0T1 | $\Delta$ T0T2 [95% CI] | Cohen's d**<br>T0T2 | P-value (T-test)<br>on<br>LLE vs IHLE<sup>1</sup> | P-value<br>(ANOVA)<br>on BMI<sup>2</sup> | P-value (T-test)<br>on<br>group size<sup>3</sup> |
| **Anthropometrics** |  |  |  |  |  |  |  |  |  |  |
| Weight | 105.63 (18.61) | 101.65 (17.59) | 101.39 (18.75) | -3.26 [-3.82; -<br>2.70]* | 0.53 | -4.13 [-4.74; -3.51]* | 0.55 | 0.07 | 0.02* | 0.97 |
| BMI | 35.97 (5.29) | 34.90 (5.40) | 34.60 (5.41) | -1.12 [-1.31; -<br>0.93]* | 0.53 | -1.40 [-1.61; -1.19]* | 0.55 | 0.15 | 0.00* | 0.84 |
| Waist<br>circumference | 116.38 (13.1) | 112.25 (14.20) | 111.72 (14.49) | -3.42 [-4.27; -<br>2.56]* | 0.42 | -4.37 [-5.17; -3.57]* | 0.51 | 0.21 | 0.12 | 0.95 |
| **Personal factors and feeling fit** |  |  |  |  |  |  |  |  |  |  |
| Self-mastery | 2.54 (0.81) | 2.44 (0.79) | 2.42 (0.73) | -0.06 [-0.13;<br>0.01] | 0.10 | -0.11 [-0.18; -0.03]* | 0.15 | 0.40 | 0.99 | 0.21 |
| Perceived<br>health | 8.93 (2.27) | 10.40 (2.08) | 10.47 (2.29) | 1.39 [1.16;<br>1.62]* | 0.58 | 1.56 [1.35; 1.77]* | 0.64 | 0.35 | 0.48 | 0.22 |
| Fitness<br>(waking) | 2.45 (1.01) | 2.68 (0.85) | 2.70 (0.89) | 0.20 [0.11;<br>0.30]* | 0.20 | 0.25 [0.17; 0.34]* | 0.25 | 0.21 | 0.76 | 0.10 |
| Construct/<br>factor | T0 M (SD) | T1 M (SD) | T2 M (SD) | $\Delta$ T0T1 [95%<br>CI] | Cohen's<br>d**<br>T0T1 | $\Delta$ T0T2 [95% CI] | Cohen's d**<br>T0T2 | P-value (T-test)<br>on<br>LLE vs IHLE<sup>1</sup> | P-value<br>(ANOVA)<br>on BMI<sup>2</sup> | P-value (T-test)<br>on<br>group size<sup>3</sup> |
| Fitness<br>(daytime) | 2.63 (0.92) | 2.70 (0.86) | 2.84 (0.85) | 0.06 [-0.04;<br>0.16] | 0.06 | 0.21 [0.12; 0.29]* | 0.22 | 0.46 | 0.88 | 0.07 |
| Support | 3.72 (1.07) | 3.72 (0.98) | 3.80 (0.94) | -0.003 [-<br>0.12;0.11] | -0.003 | 0.02 [-0.09; 1.13] | 0.02 | 0.41 | 0.81 | 1.00 |
| Influence of<br>stress on daily<br>functioning | 2.21 (0.970) | 2.27 (0.91) | 2.19 (0.89) | 0.05 [-0.04;<br>0.14] | 0.05 | -0.03 [-0.11; 0.06] | -0.03 | 0.84 | 0.39 | 0.01* |

<sup>1</sup> Comparison of two subgroups: participants with a lower level of education (LLE) to participants with an intermediate to higher level of education (IHLE).
<sup>2</sup> Comparison of different subgroups: participants with a BMI 25-30, BMI 30-35, BMI 35-40 and BMI 40+.
<sup>3</sup> Comparison of two subgroups: participants in group sizes of less than 10 participants to group sizes of 10 or more participants.
\* p<0.05
\*\* Effect size: positive values represent change in desired direction, negative values represent change in undesired direction

### Anthropometrics

The anthropometric measurements, i.e. weight, BMI and waist circumference, all showed a medium effect size in the desired direction at T1 increasing slightly at T2. Participants showed on average a decrease of 4.1 kg weight, 1.4 BMI point and 4.4 cm waist circumference after two years of CooL.

Three quarters of all participants showed weight loss during 24 months of CooL and 32% of all participants showed more than 5% weight loss. On average participants lost 3.8% weight during these 24 months.

The CooL finishers (dataset B) showed slightly better outcomes at T2, i.e. an average weight loss of 4.7 kg, a decrease of 1.6 BMI point and a decline of 5.5 cm in waist circumference at T2 (all large effect sizes).

### Personal factors and feeling fit

Participants experienced an increased feeling of self-mastery at T2 (small effect size) and an improvement in perceived health both at T1 and T2 (large effect size). Feeling fit when waking up, showed an improvement with a small effect size both at T1 and T2 whereas feeling fit during the day showed no effect at T1 and a small effect size at T2. No effect was found on perceived support and influence of stress on daily functioning both at T1 and T2 compared to baseline.

The CooL finishers (dataset B) showed similar effects and effect sizes.

All results on the behavioral factors in the full dataset (A) are summarized in Table 3 whereas more detailed data is provided in appendix II.

**Table 3.**
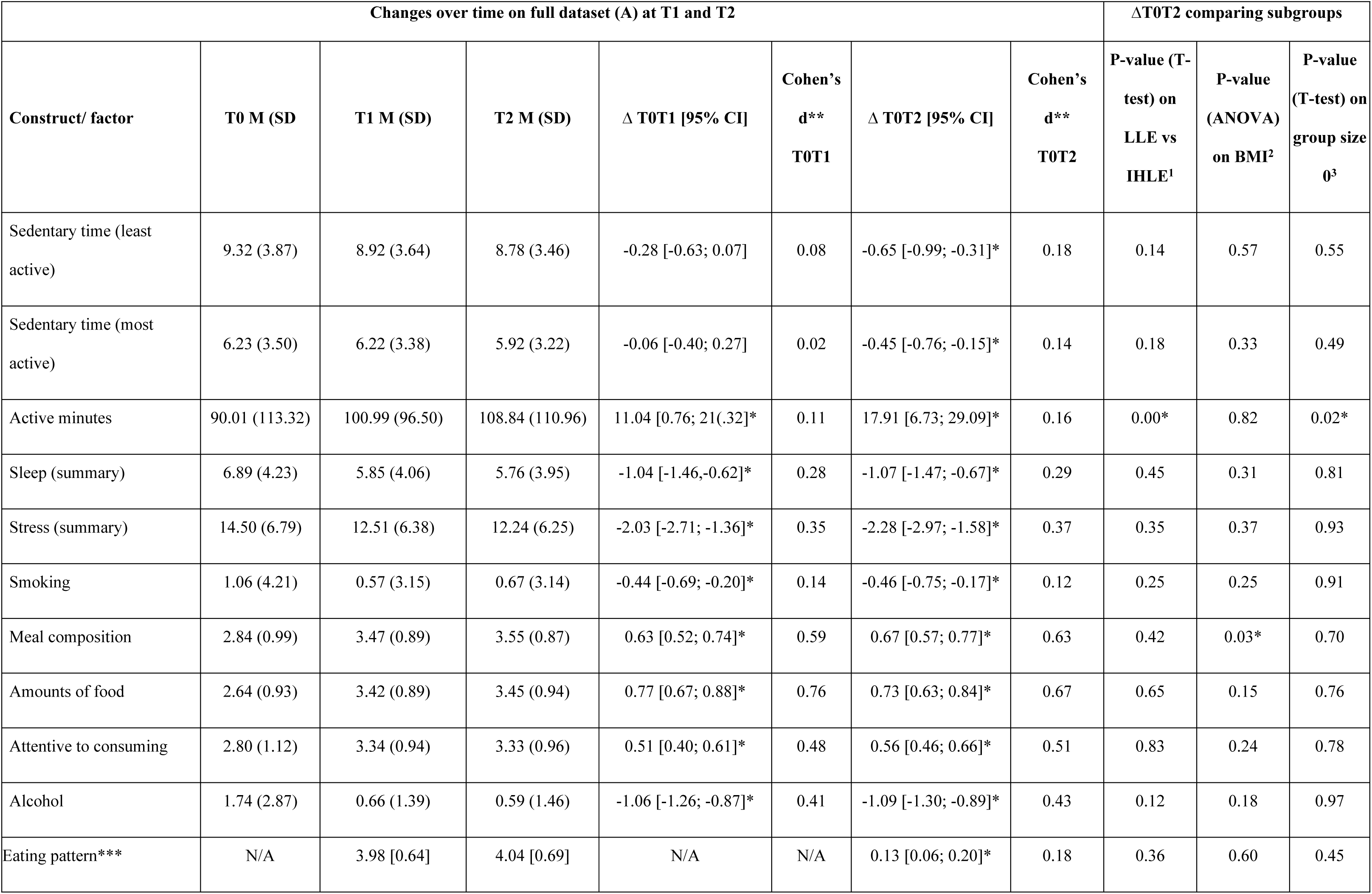
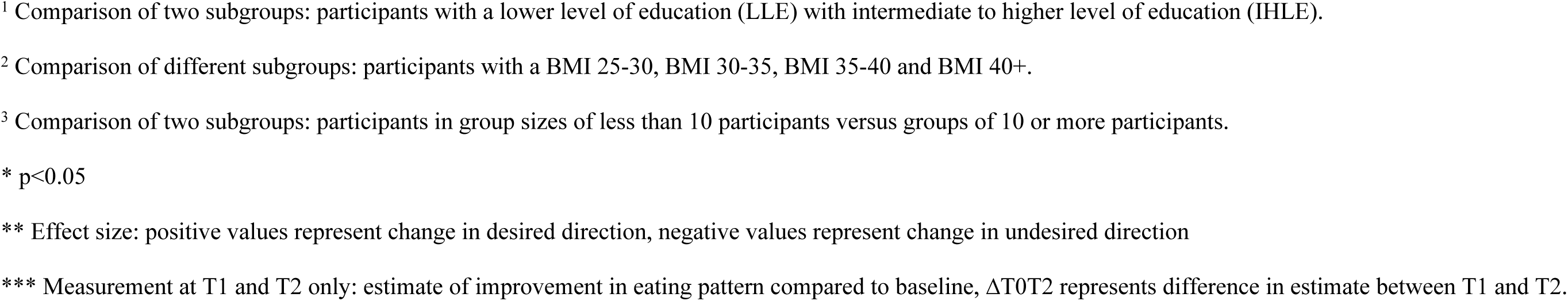
Overview of changes over time in behavioral factors in complete population and in subgroups

| Changes over time on full dataset (A) at T1 and T2 |  |  |  |  |  |  |  | ΔT0T2 comparing subgroups |  |  |
| --- | --- | --- | --- | --- | --- | --- | --- | --- | --- | --- |
| Construct/ factor | T0 M (SD) | T1 M (SD) | T2 M (SD) | Δ T0T1 [95% CI] | Cohen's<br>d**<br>T0T1 | Δ T0T2 [95% CI] | Cohen's<br>d**<br>T0T2 | P-value (T-test) on<br>LLE vs<br>IHLE<sup>1</sup> | P-value<br>(ANOVA)<br>on BMI<sup>2</sup> | P-value<br>(T-test) on<br>group size<br>0<sup>3</sup> |
| Sedentary time (least active) | 9.32 (3.87) | 8.92 (3.64) | 8.78 (3.46) | -0.28 [-0.63; 0.07] | 0.08 | -0.65 [-0.99; -0.31]* | 0.18 | 0.14 | 0.57 | 0.55 |
| Sedentary time (most active) | 6.23 (3.50) | 6.22 (3.38) | 5.92 (3.22) | -0.06 [-0.40; 0.27] | 0.02 | -0.45 [-0.76; -0.15]* | 0.14 | 0.18 | 0.33 | 0.49 |
| Active minutes | 90.01 (113.32) | 100.99 (96.50) | 108.84 (110.96) | 11.04 [0.76; 21(.32)]* | 0.11 | 17.91 [6.73; 29.09]* | 0.16 | 0.00* | 0.82 | 0.02* |
| Sleep (summary) | 6.89 (4.23) | 5.85 (4.06) | 5.76 (3.95) | -1.04 [-1.46; -0.62]* | 0.28 | -1.07 [-1.47; -0.67]* | 0.29 | 0.45 | 0.31 | 0.81 |
| Stress (summary) | 14.50 (6.79) | 12.51 (6.38) | 12.24 (6.25) | -2.03 [-2.71; -1.36]* | 0.35 | -2.28 [-2.97; -1.58]* | 0.37 | 0.35 | 0.37 | 0.93 |
| Smoking | 1.06 (4.21) | 0.57 (3.15) | 0.67 (3.14) | -0.44 [-0.69; -0.20]* | 0.14 | -0.46 [-0.75; -0.17]* | 0.12 | 0.25 | 0.25 | 0.91 |
| Meal composition | 2.84 (0.99) | 3.47 (0.89) | 3.55 (0.87) | 0.63 [0.52; 0.74]* | 0.59 | 0.67 [0.57; 0.77]* | 0.63 | 0.42 | 0.03* | 0.70 |
| Amounts of food | 2.64 (0.93) | 3.42 (0.89) | 3.45 (0.94) | 0.77 [0.67; 0.88]* | 0.76 | 0.73 [0.63; 0.84]* | 0.67 | 0.65 | 0.15 | 0.76 |
| Attentive to consuming | 2.80 (1.12) | 3.34 (0.94) | 3.33 (0.96) | 0.51 [0.40; 0.61]* | 0.48 | 0.56 [0.46; 0.66]* | 0.51 | 0.83 | 0.24 | 0.78 |
| Alcohol | 1.74 (2.87) | 0.66 (1.39) | 0.59 (1.46) | -1.06 [-1.26; -0.87]* | 0.41 | -1.09 [-1.30; -0.89]* | 0.43 | 0.12 | 0.18 | 0.97 |
| Eating pattern*** | N/A | 3.98 [0.64] | 4.04 [0.69] | N/A | N/A | 0.13 [0.06; 0.20]* | 0.18 | 0.36 | 0.60 | 0.45 |

<sup>1</sup> Comparison of two subgroups: participants with a lower level of education (LLE) with intermediate to higher level of education (IHLE).
<sup>2</sup> Comparison of different subgroups: participants with a BMI 25-30, BMI 30-35, BMI 35-40 and BMI 40+.
<sup>3</sup> Comparison of two subgroups: participants in group sizes of less than 10 participants versus groups of 10 or more participants.
\* $p < 0.05$
\*\* Effect size: positive values represent change in desired direction, negative values represent change in undesired direction
\*\*\* Measurement at T1 and T2 only: estimate of improvement in eating pattern compared to baseline, $\Delta T0T2$ represents difference in estimate between T1 and T2.

### Behavioral factors

No effect was found at T1 for sedentary time (both least and most active days) and a small effect was found at T2: participants spent around half an hour less time on sitting both on least and most active days. Physical activity showed a small effect size both at T1 and T2 with an average increase of 18 minutes at T2. The outcomes on sleep showed that participants experienced a higher quality of sleep at T1 and T2, both with a small effect size. In addition, participants experienced less stress at T1 and T2 (both medium effect size) and participants smoked less at T1 and T2 (small effect size).

The dietary outcomes showed that participants paid more attention to meal composition and to the amount of food they consume compared to baseline, both constructs showed a large effect size at T1 and T2. In addition, participants were more attentive during actual consumption of food both at T1 and T2 (both medium effect size). When drinking alcohol, participants consumed on average one unit less alcohol at T1 (small effect size) and this effect was sustained until T2 (small effect size).

Regarding change in eating pattern compared to baseline, participants indicated an improvement at T2 compared to T1 with a small effect size.

The CooL finishers (dataset B) showed deviating outcomes on physical activity (no effect at T1) and smoking (no effect at T1 or T2).

### Subgroup analyses

We compared subgroups based on the categorization of different constructs, i.e. educational level, BMI at baseline and group size at the start of the two-year CooL-intervention. All subgroup analyses were done on the full dataset (A).

#### Subgroup: educational level

Comparing participants with a lower level of education (LLE) to an intermediate or higher level of education (IHLE) showed in general no differences in effects. The only difference between both subgroups was found in active minutes. Participants with a lower level of education showed a larger increase from baseline to T2: 49 minutes of increased physical activity compared to 7 minutes for participants with a medium to higher level of education. See Tables 2 and 3 for the p-values on the subgroup comparison on educational level and appendix III for more detailed outcomes of the subgroup analysis.

#### Subgroup: BMI at baseline

For most constructs no differences in effects were observed between participants of different BMI-categories. The comparison showed differences only on change in BMI, percentage weight loss and meal composition between T0 and T2. See tables 2 and 3 for the p-values on the subgroup comparison on BMI-category and appendix IV for more detailed outcomes of the subgroup analysis. Looking into more detail, these differences were found for a limited number of categories (see appendix IV, table b).

The outcomes for participants with a larger BMI at baseline showed equal effect sizes on most constructs and better outcomes on weight loss percentage/BMI. The only exception was the attentiveness to meal composition: participants with a BMI<30 at baseline showed more improvement on this construct compared to participants with a BMI 40+.

#### Subgroup: Group size at baseline

A comparison between the outcomes of participants that start in small groups (group size <10) versus large groups (group size 10+) showed similar results. Only for two constructs differences were observed, i.e. the influence of stress on daily functioning (table 2) and the number of daily active minutes (table 3). For participants in smaller groups (<10) the influence of stress on daily functioning showed an increase at T2 compared to baseline (i.e. a more positive influence of stress on daily functioning) whereas for participants in larger groups (10+) the influence of stress showed a decrease at T2 (i.e. a more negative influence). For participants in smaller groups the physical active minutes increased on average at T2 with 38 minutes whereas for participants in larger groups the physical active minutes increased with 9 minutes compared to baseline. See appendix V for more detailed outcomes of the subgroup analysis on group sizes.

## Discussion

The results of the study showed changes in the desired direction on all outcomes at 24 months compared to baseline. The largest effect sizes were found on perceived health, attentiveness towards meal size and meal composition (large), followed by weight loss/BMI and waist circumference (medium to large). Medium effect sizes were found on attentiveness to consuming, alcohol intake and stress perception. All other behaviors showed small effect sizes whereas very few outcomes showed no effect.

Looking at changes in the timeframe of baseline to 8 months, the pattern is similar to the outcomes from previous CooL-research [8]. In addition, the present study showed sustained and improved results in CooL- participants, including enlarged weight loss over the full term of 24 months, even though they were exposed to the COVID-19 pandemic and restrictions. Durable weight loss (i.e. weight maintenance) can be defined as intentional weight loss that has been maintained for at least 6 months [27]. In general, initial weight loss is considered relatively easy whereas the opposite is true for durable weight loss. Follow-up measurements of lifestyle programs usually report weight regain compared to baseline after one year [10, 28–30] underlining the importance of the 24 months duration of CooL with a continued focus on behavior change and behavior maintenance in phase 2 of the program.

The results of the full dataset compared to the cleaned dataset were quite similar, indicating limited selective dropout. Participants that finished CooL show in broad terms slightly improved outcomes compared to the participants in the full dataset, probably because, on average, dropouts participated 11 months in CooL, whereas participants that finished CooL received additional months of guidance and support. In addition, the dropout-group consisted of participants with a range of results on both ends: participants with such positive results early in the program needing no further assistance in behavior maintenance and participants with such negative or less encouraging results wishing no longer to continue the intervention. These results on both ends of the spectrum, seem a reasonable explanation for the average result on the dropout group in total.

Despite the cut-off date that was applied to balance the dropouts in the dataset, the number of dropouts was still relatively high compared to earlier research on CooL [31]. As we suspected the COVID-19 pandemic and its restrictions to have a major influence on dropout rates, a quick analysis on the monthly dropouts from June 2019 until June 2021 showed that dropouts more than doubled when restrictions resulting from the COVID-19 pandemic came into effect starting March 2020.

Although the group of lower educated was somewhat overrepresented in the dropout group, the educational level did not seem to interfere with the achieved effects as participants with a lower level of education showed identical effects in outcomes as participants with higher educational levels, similar to the outcomes of comparable health interventions like SLIMMER [32]. Tentatively, the present study provides indications that CooL does not enlarge health inequalities and even shows potential to decrease these inequalities, under the condition that participants with a lower level of education can be guided towards sustained participation in CooL. Physical active minutes provided the only exception as participants with a lower level of education showed a larger increase despite more physical active minutes at baseline, which might be due to a confusing question on (bouts of) active minutes generating a mix of active minutes or bouts of 10 minutes.

Participants with a lower level of education were more likely to dropout during CooL which is in line with a higher dropout rate of MetSLIM, a CLI for low SES, compared to the regular SLIMMER CLI [32] and with earlier findings illustrating that higher education serves as a protective factor against dropout[33]. In conclusion, CooL is just as effective for participants with a lower level of education, though extra effort is needed to prevent dropout for this target group.

The outcomes for participants with a larger BMI at baseline showed equal effect sizes on most constructs and even better outcomes on weight loss/BMI with attentiveness to meal composition as the only exception. These outcomes indicate that the CooL-intervention is appropriate and effective for all BMI-categories.

When comparing the outcomes of participants in small groups versus large groups, the similarities stand out, as only differences were found on the impact of stress on daily functioning and on physical active minutes, both in favor of a smaller group size. However, a smaller group size is related to more dropouts, potentially due to the fact that a larger group size increases the chance of finding a suitable buddy or role model among the group as group participants and CooL-coach stay together from start to end. These findings leave the ideal group size for CooL undecided leaving room for the CooL-coach to act on personal preferences as an extra group member provides more income but requires extra effort in individual support and group dynamics.

### Limitations and strengths

A control group for comparison was no option, as the CLI CooL is part of basic health care in the Netherlands. Therefore, the results of CooL should be labelled as changes over time instead of effects as we cannot rule out interference with other factors and variables. We are less hesitant in addressing these changes to CooL given the average effect size of the changes, previous results of CooL and the comparison to similar interventions.

During the time of the study the questionnaire was revised with minor changes. We intended to keep the line of questioning and answering the same, but we cannot rule out any effect on the study. The impact of COVID-19 on the intervention, coaches and participants can be considered a second limitation though the results of the CooL-intervention on participants during COVID-19 are substantial and encouraging [8].

We cannot distinguish between dropouts and loss to follow-up as we are dependent on the respondents to hand-over their results. As a consequence, we cannot provide exact numbers on participants and dropouts of CooL. We compared the cleaned dataset with the full dataset, the latter including dropouts. The comparison is insightful but does not support firm conclusions on the (missing) effects of dropouts.

Conversely, there are several strengths to this study. This is the first study with a two-year follow-up measurement in participants of CooL: the outcomes in the long run, the nationwide inclusion and the broad scope of the research provide valuable insights on the long-term effects of CooL. In addition, the study is based on data provided by people that participate in CooL in a real-life setting. As CooL is part of basic health care insurance, it is accessible to everybody meeting the criteria and the outcomes are generalizable to those participating in real life.

### Recommendations for future research

This research provides an overview of the changes over a two-year time frame of the participants of CooL, showing more and less expected outcomes. Our recommendations for future research seize on these current outcomes:

- More research on the two-year follow-up of CLIs in the Netherlands.
- More research into the optimal group size for health interventions, in support of explicit guidelines for the healthcare workers.
- In-depth research into dropouts of the CLI, providing an overview of risk factors for dropout as well as recommendations to prevent dropout.

## Conclusion

The effects of CooL on its participants show sustained and even enlarged weight loss when comparing phase 1 to phase 2 of CooL, supporting the two-year set-up of CooL with frequent contact moments and more attention for behavioral maintenance in the second part of the intervention. The outcomes indicate that CooL is appropriate and effective for different group sizes and for a wide variety of participants irrespective of gender, age, BMI at baseline or level of education.

## Data Availability

The datasets generated and/or analyzed during the current study are not publicly available because the informed consent statement for using data was limited to the authors of this article and are only available from the corresponding author on reasonable request.

https://doi.org/10.17605/OSF.IO/Q8G7W

## Acknowledgements

The authors like to thank the CooL-coaches and the CooL-participants for their efforts to make this research possible.

## Supporting information

**S1 Table. Predictors for dropout.** Logistic regression outcomes on predictors for dropout.

**S2 Table. Changes over time CooL finishers.** Detailed overview on changes over time on participants that completely finished CooL.

**S3 Table. Subgroup comparison by educational level.** Detailed overview of the subgroup comparison on educational level.

**S4 Table. Subgroup comparison by BMI category.** Detailed overview of the subgroup comparison on BMI category.

**S5 Table. Subgroup comparison by group size.** Detailed overview of the subgroup comparison of larger group size versus smaller group size.

